# Repeat COVID-19 Molecular Testing: Correlation with Recovery of Infectious Virus, Molecular Assay Cycle Thresholds, and Analytical Sensitivity

**DOI:** 10.1101/2020.08.05.20168963

**Authors:** Victoria Gniazdowski, C. Paul Morris, Shirlee Wohl, Thomas Mehoke, Srividya Ramakrishnan, Peter Thielen, Harrison Powell, Brendan Smith, Derek T. Armstrong, Monica Herrera, Carolyn Reifsnyder, Maria Sevdali, Karen C. Carroll, Andrew Pekosz, Heba H. Mostafa

**Author notes:** Authors contributed equally (Victoria Gniazdowski, C. Paul Morris).

## Abstract

Repeat molecular testing for SARS-CoV-2 may result in scenarios including multiple positive results, positive test results after negative tests, and repeated false negative results in symptomatic individuals. Consecutively collected specimens from a retrospective cohort of COVID-19 patients at the Johns Hopkins Hospital were assessed for RNA and infectious virus shedding. Whole genome sequencing confirmed the virus genotype in patients with prolonged viral RNA shedding and droplet digital PCR (ddPCR) was used to assess the rate of false negative standard of care PCR results. Recovery of infectious virus was associated with Ct values of 18.8 ± 3.4. Prolonged viral RNA shedding was associated with recovery of infectious virus in specimens collected up to 20 days after the first positive result in patients who were symptomatic at the time of specimen collection. The use of Ct values and clinical symptoms provides a more accurate assessment of the potential for infectious virus shedding.

## Introduction

Molecular methods for SARS-CoV-2 nucleic acid detection from nasopharyngeal swabs have been the gold standard for COVID-19 diagnosis. Although diagnostic approaches target different genes within the SARS-CoV-2 genome, they have shown comparable analytical sensitivity and high specificity (1–17). Sensitivity of the assay is associated with the shedding pattern of SARS-CoV-2 RNA, which can vary based on the source of respiratory specimen and based on the course of illness (18–21).

Infection control personnel and physicians managing COVID-19 patients and patients under investigation (PUI) continue to face several diagnostic dilemmas related to a lack of understanding of the clinical sensitivities of SARS-CoV-2 molecular diagnostics and the correlation between viral RNA detection and shedding of infectious virus. Retesting of patients has become a common practice especially when there is a strong clinical suspicion or exposure history and there is an initial negative result (22). A single positive molecular result should be sufficient for confirming COVID-19 diagnosis, however, repeated testing of hospitalized patients for determining isolation needs and infection control measures has become a part of managing this patient population. Two negative molecular assay results from two consecutively collected respiratory specimens more than 24 hours apart has been the strategy used for discontinuation of transmission precautions and returning to work (23). Repeat testing on patients has revealed that SARS-CoV-2 RNA can be detectable for weeks after the onset of symptoms (24). In addition, there have been reports of patients who had initial negative molecular tests that tested positive on subsequent tests. In general, molecular detection of SARS-CoV-2 RNA does not necessarily denote the presence of recoverable infectious virus. A few studies, as well as data from the CDC, showed that higher viral loads are associated with recovery of infectious virus and that virus recovery is generally not reported after 9 days from symptom onset (20, 25, 26). A case study, in which severe infection was associated with recovery of infectious SARS-CoV-2 from stool indicates that the duration of recovery of infectious virus particles might vary based on the severity of the disease or the duration of symptoms (27).

False negative molecular SARS-CoV-2 results occur and in some cases a single negative result is not sufficient for excluding COVID-19 diagnosis. False negative rates are estimated to range from 5 to 40%, yet a conclusive percentage is currently difficult to determine due to the lack of a diagnostic comparator gold standard (28, 29). Initial false negative results in the setting of consistent respiratory symptoms have been reported, with some patients having subsequent positive results on serial testing (30). The Infectious Diseases Society of America (IDSA) recommends repeated testing after initial negative RNA testing in cases with intermediate to high suspicion of COVID-19, but evidence that this practice positively affects outcomes is still lacking (31). Clinical sensitivity has also been attributed to the specimen type collected and the time of collection in relation to the duration of symptoms (32–42).

In this study, we analyzed the molecular diagnostics data from Johns Hopkins Hospital in the time frame March 11^th^ to May 11^th^ 2020. Our study aimed to dissect different diagnostic dilemmas by incorporating statistics of repeat testing, cycle threshold values, infectious virus isolation, whole genome sequencing, and ddPCR. We address questions that include: 1) How does a positive molecular test correlate with recovery of infectious virus? 2) Are patients with prolonged viral RNA shedding also shedding infectious virus? 3) Are there changes in viral sequences during prolonged shedding? 4) Does a positive test result following undetectable viral RNA correlate with infectious virus recovery? 5) Can false negative results due to an assay’s analytical limitation (limit of detection) be detected by ddPCR?

## Methods

### Study site and ethics

This study was performed in the Molecular Virology Laboratory, Johns Hopkins Hospital. Cell culture studies were conducted at the Johns Hopkins Bloomberg School of Public Health. The study was approved by the Johns Hopkins University School of Medicine Institutional Review Board. The aggregate metadata of the selected patient population for further studies is shown in supplementary table 1.

### Clinical data, standard of care assays, and specimens

Repeat testing was identified by pulling the data of all molecular COVID-19 testing that was conducted in the Johns Hopkins Hospital Microbiology laboratory from March 11^th^ to May 11^th^ 2019. Data were pulled using the laboratory information system (Soft). Specimens used were remnant specimens available at the completion of standard of care testing at the Johns Hopkins Laboratory. During the time frame reported, several molecular diagnostic assays for SARS-CoV-2 were used including The RealStar® SARS-CoV-2 RT-PCR Kit 1.0 from Altona Diagnostics (Hamburg, Germany) (3), the CDC COVID-19 RT-PCR panel assay, the GenMark (Carlsbad, CA) ePlex® SARS-CoV-2 Test (3, 43), the NeuModx™ SARS-CoV-2 Assay (44), the BD SARS-CoV-2 Reagents For BD MAX™ System (45), and the Xpert Xpress SARS-Cov-2 (46). The Ct values shown are for specimens diagnosed by either the RealStar® or the NeuModx™ SARS-CoV-2 assays. For simplicity, we show the Ct values of only one gene target per assay: the Spike (S) gene for the RealStar® SARS-CoV-2 and the nonstructural protein (Nsp) 2 gene for the NeuMoDx™ SARS-CoV-2 assays. Our data indicates comparable Ct values for the two genes (Mostafa *et al*, under revision).

### Nucleic acid extractions

Nucleic acid extractions for the RealStar® SARS-CoV-2, the CDC COVID-19 RT-PCR panel, the ddPCR assays, and Nanopore whole genome sequencing were performed as previously described in (3). The NucliSENS easyMag or eMAG instruments (bioMérieux, Marcy-l’Etoile, France) were used using software version 2.1.0.1. The input specimens’ volumes were 500 μL and the final elution volume was 50 μL. Specimens for automated systems were processed following each assay’s FDA-EUA package insert.

### SARS-CoV-2 Virus Isolation

VeroE6 cells (ATCC CRL-1586) were cultured at 37°C with 5% carbon dioxide in a humidified chamber using complete medium (CM) consisting of Dulbecco’s modified Eagle Medium supplemented with 10% fetal bovine serum (Gibco), 1mM glutamine (Invitrogen), 1mM sodium pyruvate (Invitrogen), 100μg/mL penicillin (Invitrogen) and 100 μg/mL streptomycin (Invitrogen). Cells were plated in 24 well dishes and grown to 75% confluence. The CM was removed and replaced with 150 μL of infection media (IM) which is identical to CM but with the fetal bovine serum reduced to 2.5%. Fifty μL of the clinical specimen was added to one well and the cells incubated at 37°C for one hour. The inoculum was aspirated and replaced with 0.5 ml IM and the cells cultured at 37°C for 4 days. When cytopathic effect was visible in most of the cells, the IM was harvested and stored at −70°C. The presence of SARS-CoV-2 was verified by one of two ways. SARS-CoV-2 viral RNA was extracted using the Qiagen Viral RNA extraction kit (Qiagen) and viral RNA detected using quantitative, reverse transcriptase PCR (qPCR) as described (47). SARS-CoV-2 viral antigen was detected by infecting VeroE6 cells grown on 4 chamber LabTek slides (Sigma Aldrich) with 50 μL of virus isolate diluted in 150 μL of IM for 1 hour at 37°C. The inoculum was replaced with IM and the culture incubated at 37°C for 12-18 hours. The cultures were fixed with 4% paraformaldehyde for 20 minutes at room temperature and processed for indirect immunofluorescence microscopy as described (48). The humanized monoclonal antibody D-006 (Sino Biological) was used as the primary antibody to detect Spike or S protein, followed by Alexa Fluor 488-conjugated goat anti-human IgG. The cells were mounted on Prolong antifade and imaged at 40X on a Zeiss Axio Imager M2 wide-field fluorescence microscope (49).

### Oxford Nanopore whole genome sequencing

Whole genome sequencing was conducted using the Oxford Nanopore platform following the ARTIC protocol for SARS-CoV-2 sequencing with the V3 primer set (50). Eleven indexed samples (and one negative control) were pooled for each sequencing run and 20 ng of the final pooled library was run on the Oxford Nanopore GridION instrument with R9.4.1 flowcells. Basecalling and demultiplexing was performed with Guppy v3.5.2 and reads were assembled using a custom pipeline modified from the ARTIC network bioinformatics pipeline (https://artic.network/ncov-2019). As part of this custom pipeline, reads were mapped to a SARS-CoV-2 reference genome (GenBank MN908947.3) using minimap2 (51). Coverage was normalized across the genome and variant calling was performed with Nanopolish v0.13.2 (52). Sites with low coverage (based on the negative control coverage) were masked as ‘N’. Variant calls were also independently validated with two other variant callers—medaka (https://nanoporetech.github.io/medaka/snp.html) and samtools(https://wikis.utexas.edu/display/bioiteam/Variant+calling+using+SAMtools)—and all sites with disagreements or allele frequency <75% were manually inspected in Integrated Genome Viewer (53). Sites with minor allele frequency 25-75% were replaced with IUPAC ambiguity codes.

### Reverse Transcription Droplet Digital PCR (ddPCR)

The ddPCR procedure followed the assay’s EUA package insert (54). Briefly, RNA isolated from NP specimens (5.5 μL) were added to the mastermix comprised of 1.1 μL of 2019-nCoV CDC ddPCR triplex assay, 2.2 μL of reverse transcriptase, 5.5 μL of supermix, 1.1 μL of Dithiothreitol (DTT) and 6.6 μL of nuclease-free water. Twenty-two microliters from these samples and mastermix RT-ddPCR mixtures were loaded into the wells of a 96-well PCR plate (Bio-Rad, Pleasanton, CA). The mixtures were then fractionated in up to 20,000 nanoliter-sized droplets in the form of a water-in-oil emulsion in the Automated Droplet Generator (Bio-Rad, Pleasanton, CA). The 96-well RT-ddPCR ready plate containing droplets was sealed with foil using a plate sealer (Bio-Rad, Pleasanton, CA) and thermocycled to achieve reverse transcription of RNA followed by PCR amplification of cDNA in a C1000 Touch thermocycler (Bio-Rad, Pleasanton, CA). Following PCR, the plate was loaded into the QX200 Droplet Reader (Bio-Rad, Pleasanton, CA); the droplets in each well were singulated and flowed past a two-color fluorescence detector. The fluorescence intensity of each droplet was measured in FAM and HEX, and droplets were determined to be positive or negative for each target within the Bio-Rad SARS-CoV-2 ddPCR Test: N1, N2 and RP. The fluorescence data was then analyzed by QuantaSoft 1.7 and QuantaSoft Analysis Pro 1.0 Software to determine the presence of SARS-CoV-2 N1 and N2 in the specimen.

## Results

### COVID-19 testing in the Johns Hopkins Hospital Network

The Johns Hopkins molecular virology laboratory processed a total of 29,687 COVID-19 molecular diagnostic tests from 16,968 patients (or patients under investigation) from March 11^th^ 2020 (first day of in house testing) to May 11^th^ 2020. There were 2,194 patients tested more than once with 1,788 patients repeatedly testing negative. 132 patients continued to have positive results in all the time points tested while 124 patients had an initial negative result that was followed by a positive result. 150 patients had an initial positive result that was followed by a negative test (figure 1A and B). Our data indicates that of all the patients that had repeat testing, 81.5% continued to have negative results, 5.7% had an initial negative followed by a repeat positive test, and 6.8% had a final negative test result after an initial positive (figure 1B).

**Figure 1.**
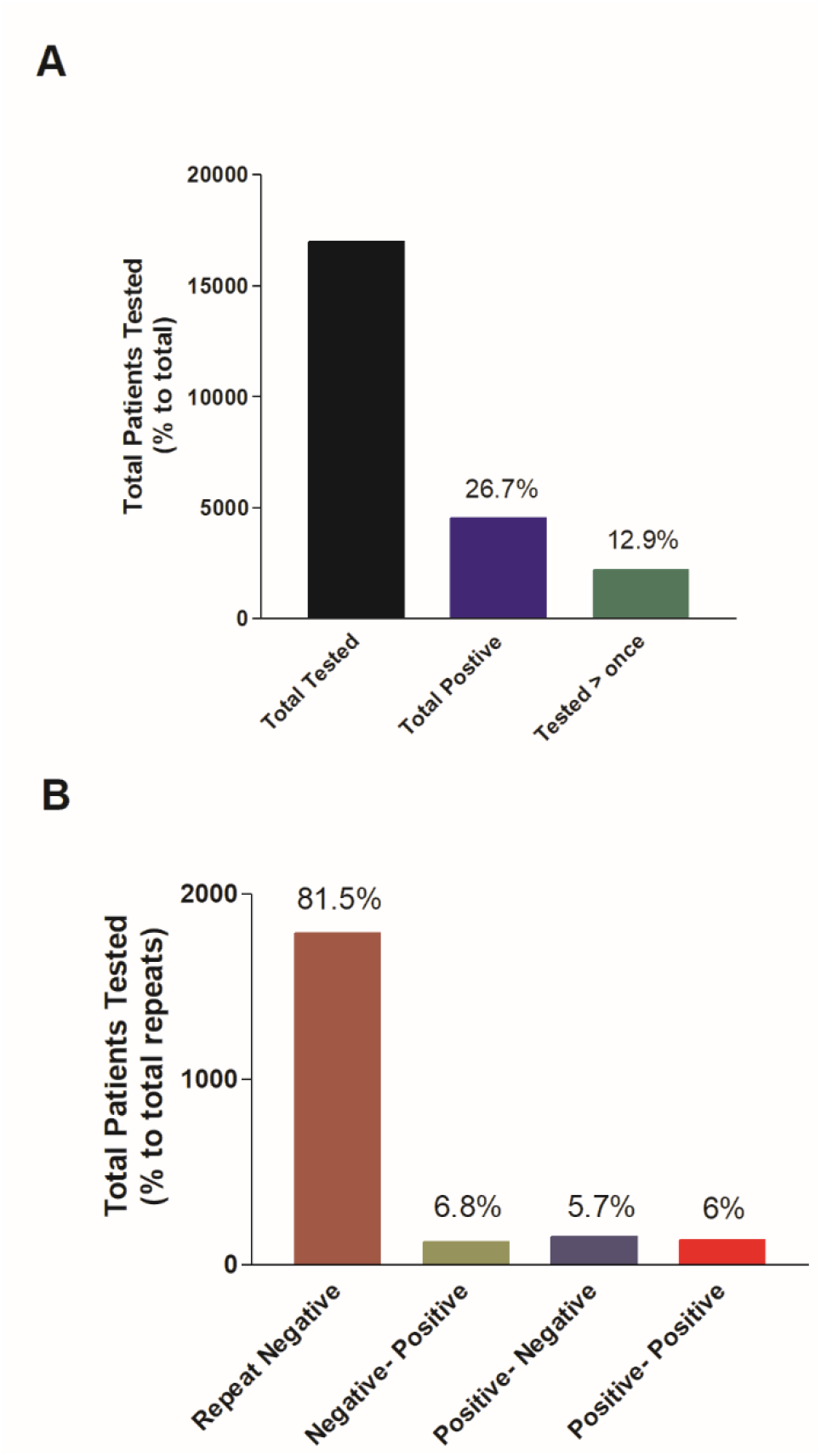
COVID-19 molecular testing at the Johns Hopkins Hospital. A) The total number of patients tested from March 11^th^ through May 11^th^ 2020, total positives, and patients tested more than once. B) Total number of patients who received repeat testing distributed based on the consecutive assays’ results.

### Infectious virus isolation and viral RNA load

To understand the correlation between a positive molecular result and virus recovery, 161 patients’ specimens that were positive by molecular testing were cultured on VeroE6 cells. The cultured specimens spanned a wide range of cycle threshold values reflecting different viral loads. The recovery of virus and the development of cytopathic effect were monitored for up to 4 days post infection of VeroE6 cells. The mean and median Ct values associated with recoverable virus were 18.8 ± 3.4 and 18.17 respectively, which was significantly lower than the mean and median Ct values that did not correlate with infectious virus recovery (27.1 ± 5.7 and 27.5 respectively) (paired t test, P<0.0001) (Figure 2). Samples with a Ct value below 23 yielded 91.5% of virus isolates. However, 28.6% of specimens that were negative for viral growth on VeroE6 cells were in that same Ct value range (Figure 2) and 11.9% were below a Ct value of 20.

### Prolonged viral RNA detection and infectious virus load

Patients that received repeated testing with longitudinal positive results were tested within a time frame that ranged from less than one day to more than 45 days. To assess the correlation between the repeated positivity, viral loads, and recovery of infectious virus, we evaluated a randomly selected subset of 29 patients. We examined the Ct values of all test results, days between testing, as well as viral growth on cell culture (if performed) (Table 1). Except for two patients (#24 and 25) (and the first three whose clinical information was not accessible), this cohort of patients had chronic underlying conditions. The observed general trend was an increase in the Ct values over time indicating a reduction in the viral RNA load, and further correlating, in the majority of the patients, with failure to recover infectious virus on cell culture. Interestingly, 4 patients had infectious virus recovered from specimens collected in up to 22 days after the first positive result, however, infectious virus shedding was not associated with a specific outcome as one patient was never admitted (# 24), one was hospitalized with no oxygen requirements (# 10), and two had more severe disease (#8 and #29). Recovery of infectious virus was associated with persistence of symptoms in all but one patient (# 24). Longitudinal specimens of patients were sequenced to assess any changes in the viral genome that could have resulted in prolonged shedding or could possibly suggest a reinfection. The successful recovery of complete viral genome sequences at multiple time points from 7 patients provided evidence that these patients were carrying the same virus over time, however in one case, the second time-point sample had additional variants, and in two cases minor variants appear in the later time point sample (denoted as IUPAC ambiguity codes, since two alleles are present in the sequencing reads) (Table 2). Of note, two different isolates collected from patient #14 in the same day were included in this analysis for validating our sequencing reproducibility.

**Figure 2.**
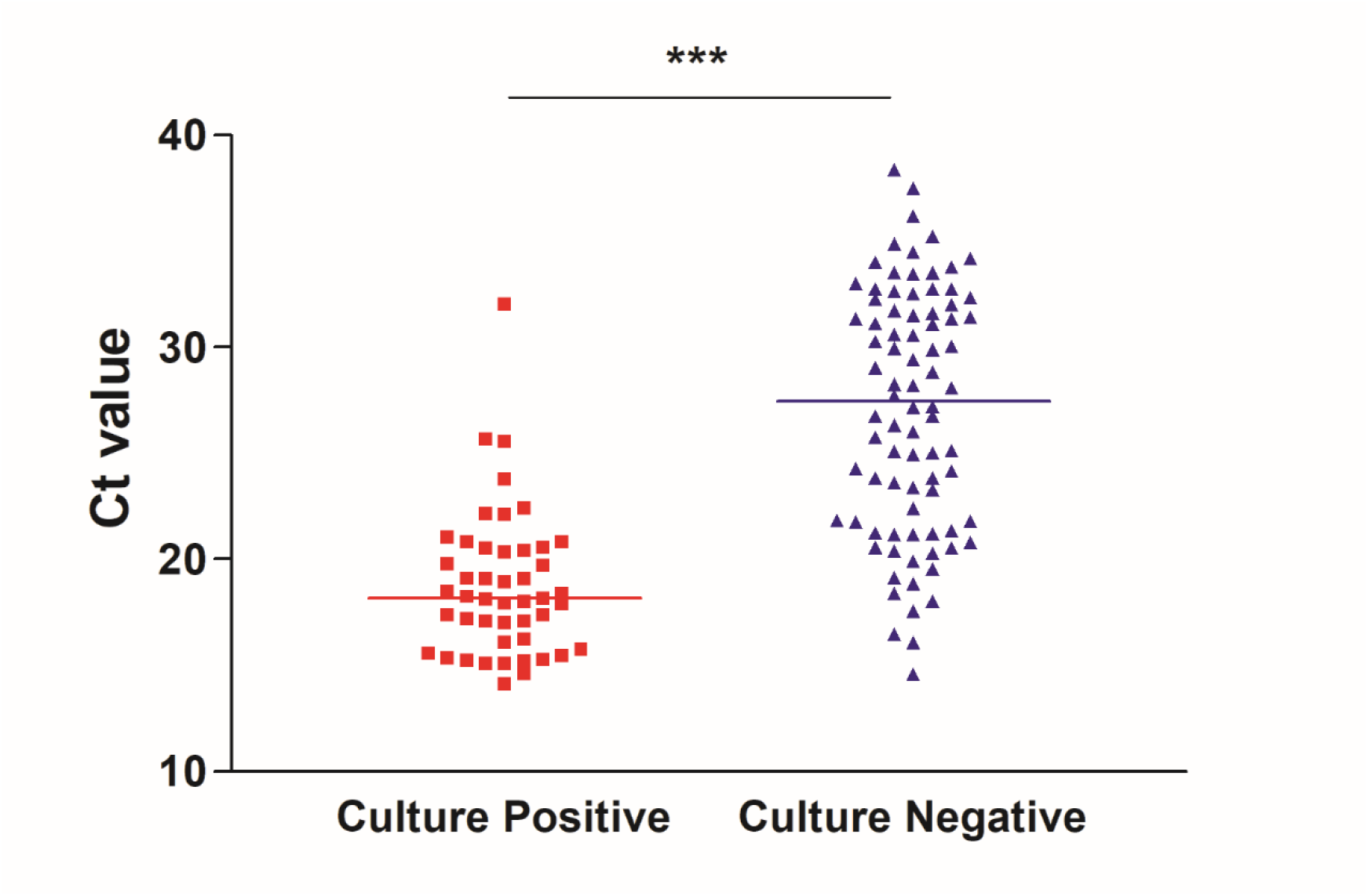
Correlation between recovery of SARS-CoV-2 infectious virus on cell culture and Ct values. Nasopharyngeal specimens were cultured on VeroE6 cells and the recovery of virus and the development of cytopathic effect were monitored for up to 4 days post infection. Viral growth was confirmed by antigen staining or PCR. *** paired *t* test, P<0.0001

**Table 1.**
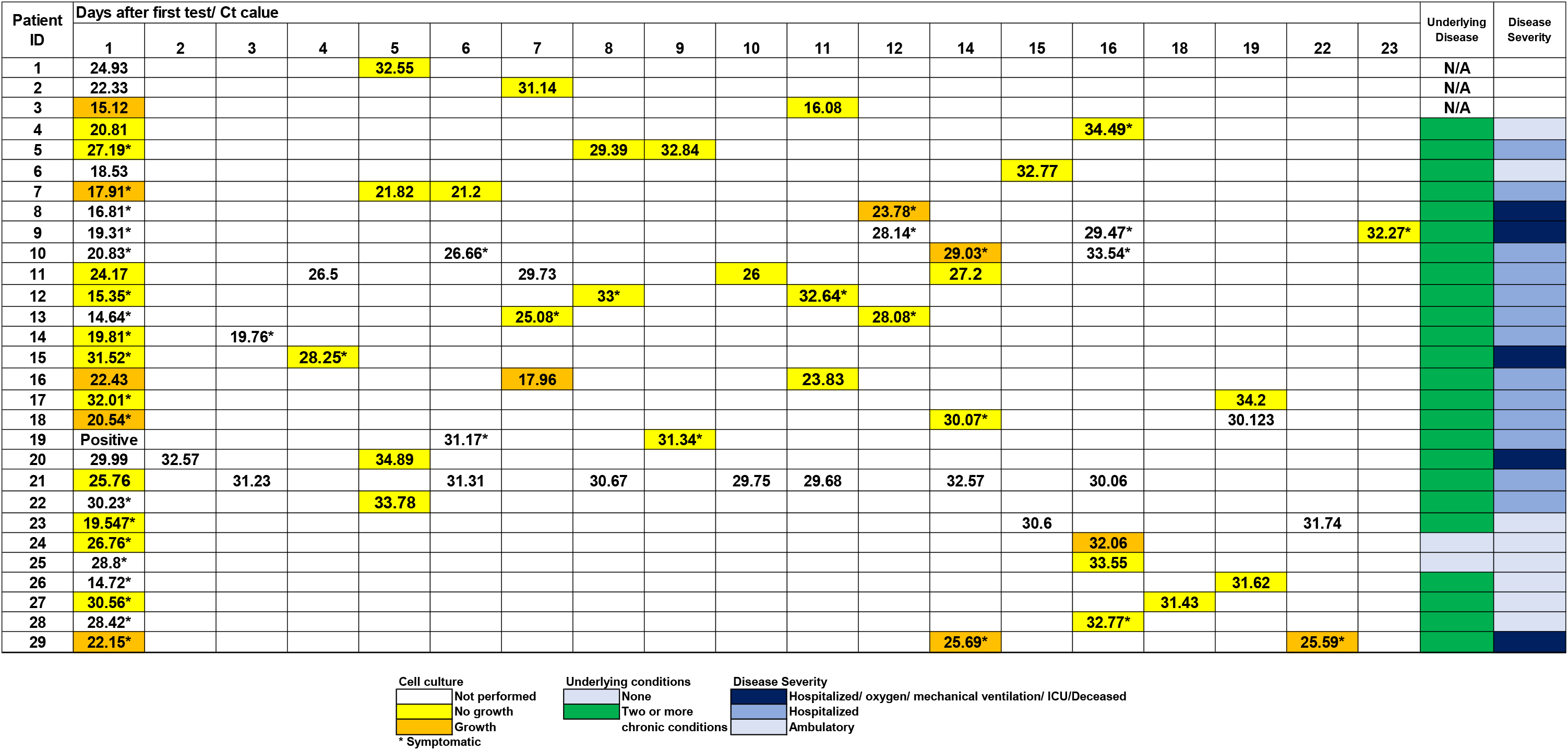
Patients with multiple positive molecular results overtime and correlation between the time of testing, isolation of infectious virus on cell culture, and the cycle threshold (Ct) value of the diagnostic assay. *symptomatic at the time of specimen collection. N/A: Not Available

**Table 2.** Sequence comparison of whole viral genomes from consecutive positive NP samples (subset of patients from table 1).

| Patient ID | Days after first test | Variants present in all as compared to Severe acute respiratory syndrome coronavirus 2 isolate Wuhan-Hu-1 | Additional variants | GISAID ID | Nextstrain clade | Nextstrain parent clade | Pangolin clade |
| --- | --- | --- | --- | --- | --- | --- | --- |
| 5 | D1 | C241T, C1059T, C3037T, C12412T, C14408T, G15760A, A23403G, G25563T |  | hCoV-19/USA/DC-HP00462/2020 | 20C | 20A | B.1 |
|  | D8 | C241T, C1059T, C3037T*, C12412T, C14408T, G15760A, A23403G*, G25563T | T4075Y, G4076K, C14456T, T20310Y, C23591T, C26533Y, C28445Y | hCoV-19/USA/DC-HP00799/2020 | -- |  | B.1 |
| 9 | D1 | C241T, C1059T, C3037T, C14408T, A23403G, G25563T |  | hCoV-19/USA/MD-HP00028/2020 | 20C | 20A | B.1 |
|  | D11 | C241T, C1059T, C3037T, C14408T, A23403G, G25563T |  | hCoV-19/USA/MD-HP00554/2020 | 20C | 20A | B.1 |
| 10 | D1 | C241T, C3037T, T9172C, A10948G, C14408T, A23403G, G26730T, C27874T, G28881A, G28882A, G28883C |  | hCoV-19/USA/MD-HP00160/2020 | 20B | 20A | B.1.1.9 |
|  | D6 | C241T, C3037T, T9172C, A10948G, C14408T, A23403G, G26730T, C27874T, G28881A, G28882A, G28883C |  | hCoV-19/USA/MD-HP00377/2020 | 20B | 20A | B.1.1.9 |
|  | D14 | C241T, C3037T, T9172C, A10948G, C14408T, A23403G, G26730T, C27874T, G28881A, G28882A, G28883C |  | hCoV-19/USA/MD-HP00885/2020 | 20B | 20A | B.1.1.9 |
| 13 | D1 | T490A, C3177T, C6040T, C6449T, C8782T, G12478A, T17531C, T18736C, C24034T, T26729C, G28077C, T28144C, C28896G, A29700G |  | hCoV-19/USA/MD-HP00567/2020 | 19B |  | A.3 |
|  | D7 | T490A, C3177T, C6040T, C6449T, C8782T, G12478A, T17531C, T18736C, C24034T, T26729C, G28077C, T28144C, C28896G, A29700G* |  | hCoV-19/USA/MD-HP00883/2020 | 19B |  | A.3 |
| 14 | D1 | C241T, C1059T, C3037T, G5555A, C14408T, A23403G, G24368C, G25563T, C27005T |  | hCoV-19/USA/MD-HP01661/2020 | 20C | 20A | B.1 |
|  | D1 | C241T, C1059T, C3037T, G5555A, C14408T, A23403G, G24368C, G25563T, C27005T |  | hCoV-19/USA/MD-HP01656/2020 | 20C | 20A | B.1 |
| 16 | D1 | T490A, C3177T, C6040T, C8782T, C8950T, G12478A, T18736C, C24034T, T26729C, G28077C, T28144C, C28896G, C29451T, A29700G |  | hCoV-19/USA/MD-HP00171/2020 | 19B |  | A.3 |
|  | D6 | T490A, C3177T, C6040T, C8782T, C8950T, G12478A, T18736C, C24034T, T26729C, G28077C, T28144C, C28896G, C29451T, A29700G |  | hCoV-19/USA/MD-HP00549/2020 | 19B |  | A.3 |
| 18 | D1 | T490A, C3177T, C6040T, C8782T, G12478A, T18736C, C24034T, T26729C, G28077C, T28144C, C28896G, A29700G | C19488Y | hCoV-19/USA/MD-HP00031/2020 | 19B |  | A.3 |
|  | D14 | T490A, C3177T, C6040T, C8782T, G12478A, T18736C, C24034T, T26729C, G28077C, T28144C, C28896G, A29700G** | C8262M, A10859W, C11844Y | hCoV-19/USA/MD-HP00336/2020 | 19B |  | A.3 |
| 29 | D1 | C241T, C1059T, C3037T, C3141A, A4919G, C14408T, A23403G, G25563T, C26625T |  | hCoV-19/USA/MD-HP02026/2020 | 20C | 20A | B.1 |
|  | D14 | C241T, C1059T, C3037T, C3141A, A4919G, C14408T, A23403G, G25563T, C26625T |  | hCoV-19/USA/MD-HP02027/2020 | 20C | 20A | B.1 |

\* Limited read data is consistent with specified mutation (>75% of reads support variant), but position is ambiguous (N) due to low coverage
\*\* Limited read data provides some evidence for possible mutation or mixture (<75% of reads support variant), but position is ambiguous (N) due to low coverage

### Testing based discontinuation of transmission precautions for COVID-19 patients

Many patients who tested negative for SARS-COV-2 showed a subsequent positive result. A subset of patients who received repeated testing and had mixed negative and positive results were examined for the Ct values of the positives that follow negative results as well as the recovery of infectious virus. The follow up positive testing on previously negative patients produced Ct values higher than 29.5 (Table 3). Attempted recovery of infectious virus from these specimens was negative.

**Table 3.**
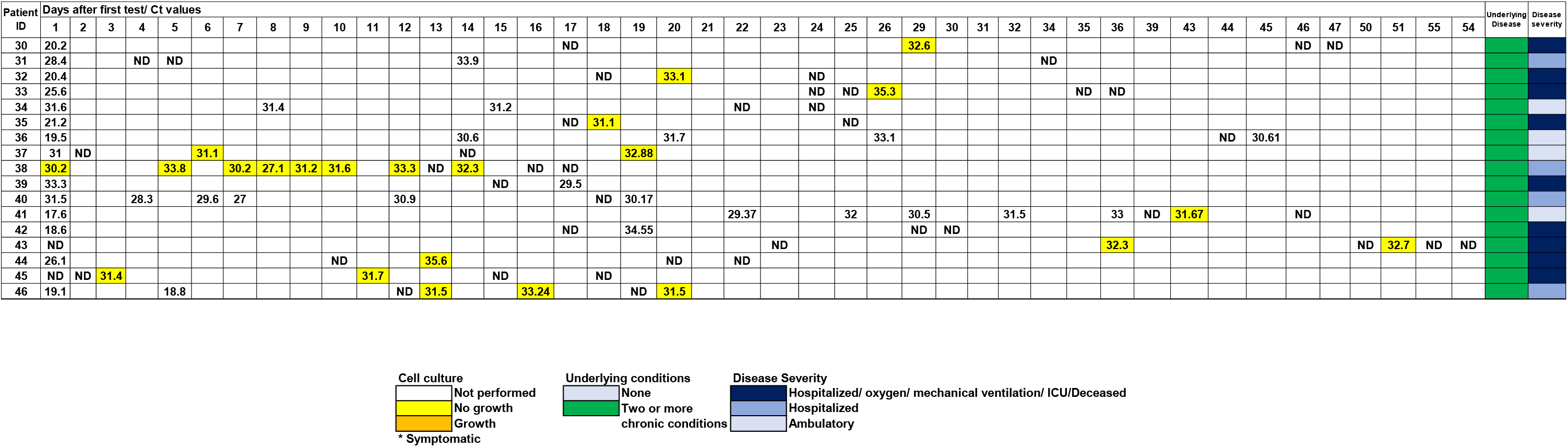
Patients with positive molecular results after one or more negatives and correlation with the time of testing, isolation of infectious virus on cell culture, and the cycle threshold (Ct) value of the diagnostic assay. ND, target not detected.

### Repeat negative testing of patients with clinical disease or exposure history with COVID-19

1,788 patients were tested more than once between March 11^th^ and May 11^th^ 2020 without any positive result. To examine the possibility of false negative results of the standard of care molecular SARS-CoV-2 diagnostic assay due to limitations in the analytical sensitivity, we used the SARS-CoV-2 droplet digital PCR (ddPCR). We selected 198 negative from 185 patients that received repeated testing over time, of which 163 patients had from 2 to up to 5 negative results. We selected 15 that had positive SARS-CoV-2 serology and multiple negative RT-PCR results. A few included 22 specimens from patients who had an initial positive result but turned negative on a repeat test or the reverse. Of the total 198 tested, 11 specimens were positive by ddPCR (Table 4). Only one patient who had a positive serology test (patient # 51) had a positive ddPCR result and 4 of the 11 patients had positive specimens by RT-PCR collected at other days (54–57).

**Table 4.**
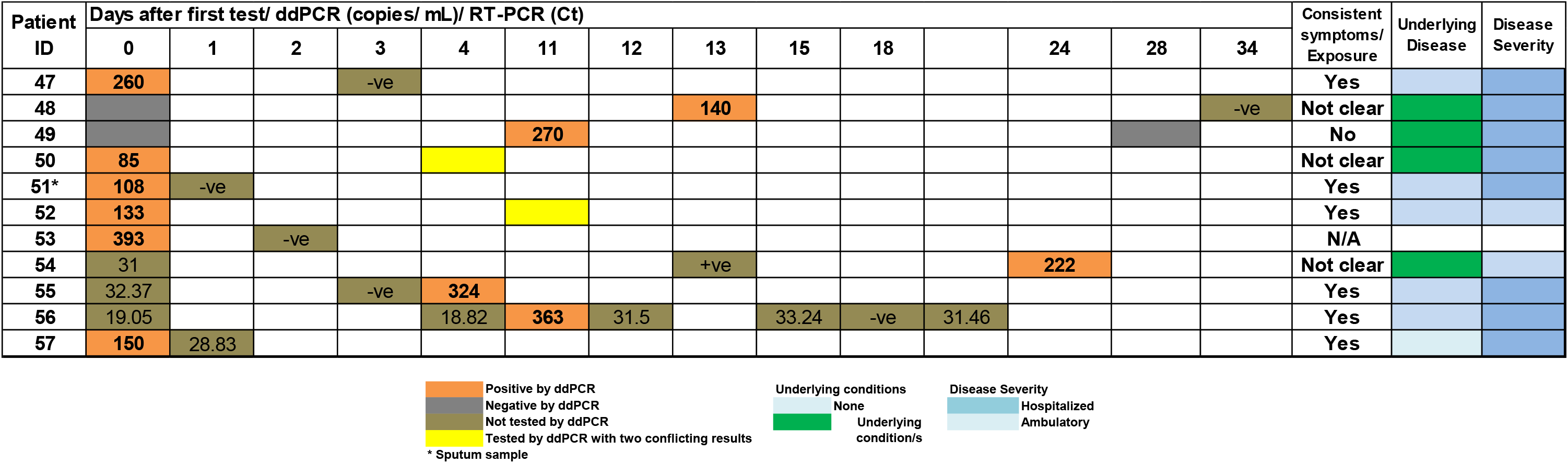
ddPCR sensitivity of detection in patients with consecutive negative results (47–53) and negative specimens collected from known positive patients (54–57). ddPCR copies shown for the N1 target. −ve: negative result by the standard of care RT-PCR. +ve: positive results by the standard of care RT-PCR with no available Ct value.

## Discussion

The molecular detection of SARS-CoV-2 genome has been valuable not only in diagnosis, but also in making decisions related to infection control measures and return to work. Several outcomes were observed with repeat molecular testing including: i) prolonged, consistent viral RNA shedding, ii) alternating negative results and positive RNA shedding, and iii) false negative results. Our data shows that prolonged positivity could be associated with recovery of infectious virus especially when symptoms persist. Our data also shows that RNA positive specimens after a negative result are not associated with recovery of infectious virus.

The ddPCR assay detected a few positives that were missed by our standard of care testing in the subset of patients who were highly suspected of infection based on clinical symptoms. Overall, our data confirms that SARS-CoV-2 RNA is detectable for a prolonged time, and recovery of infectious virus is associated with persistent symptoms. Importantly, our data also shows that the standard of care molecular diagnostics’ analytical sensitivities are affected by the shedding pattern of the viral RNA rather than the assay’s performance.

The use of a diagnostic test’s Ct values as an indicator of the presence of infectious virus has been proposed. One report suggested that a Ct value above 33-34 is not associated with recovery of infectious virus (55) and another report concluded that cell culture infectivity is observed when the Ct values were below 24 and within 8 days from symptoms onset (25). Our data shows that the average Ct value that was associated with cell culture growth is 18.8. Recovery of infectious virus was possible from some specimens with Ct values as high as 32.1 and in others that were collected up to 22 days after the first positive result, especially in patients symptomatic at the time of sample collection. A recent report showed recovery of infectious virus for a prolonged time in severely ill COVID-19 patients which could correlate with high Ct values (56). This indicates that neither the Ct values nor cell culture results should be used to make clinical decisions, or infection control decisions, due to the lack of sufficient clinical outcome studies.

A significant number of our cultured specimens that yielded no infectious virus had low Ct values (28.6% Ct < 23, figure 1) indicating that variables other than the viral genome copies play a role in isolating infectious virus on cell culture. The integrity of the viral genome and variables related to sampling and storage of specimens have been proposed to impact infectious virus recovery (57). Virus particles may be bound to neutralizing antibodies and therefore unable to initiate infection (58). Generally, prolonged shedding of viral RNA was previously noted for many other viruses, including SARS-CoV, MERS-CoV, influenza, and measles viruses (59–63).

Positive molecular results after negative tests were noticed in patients with COVID-19 and it is not certain if that indicates a relapsed infection or reinfection. Our data showed that positive RNA results detected after viral clearance (undetectable viral RNA) were not associated with recovery of infectious virus. It is likely that detectable viral RNA in convalescence is associated with prolonged viral RNA shedding especially since the viral loads are usually lower than that detectable during the early stages of infection. In addition, positive test results after negative molecular RNA tests that are associated with new symptoms are more perplexing, and reinfection has not been ruled out. Comprehensive studies that combine understanding the development of protective immunity and compare isolated viral genomes will help understanding the enigma of reinfection by SARS-CoV-2.

DdPCR showed a slightly higher sensitivity in detecting SARS-CoV-2 RNA in a subset of specimens from patients with high suspicion of COVID-19 and negative standard RT-PCR. Our data is consistent with published reports that compared ddPCR with real-time PCR (33). It is important to note that the analytical sensitivity of the ddPCR assay as reported by the EUA package insert (645 copies/ mL) is comparable to standard of care real-time PCR methods we use in our diagnostic laboratories that include the CDC panel assay among others (3) and all of the positives detected by the ddPCR assay in this study were below the ddPCR assay’s analytical limit of detection (Table 4). The Bio-Rad ddPCR assay uses primers and probes that are same as reported by the CDC assay and also includes the human RNase P gene as an internal control. Including this control is very valuable to exclude insufficient sampling as a cause of false negative results (64). Only a few samples that tested negative by the standard PCR methodologies were later positive by ddPCR (5.7%), even in a cohort with a high suspicion of COVID-19. A few samples showed conflicting results when repeated (Table 4), likely because of viral loads below the lower limit of detection of the ddPCR assay. Overall, this suggests that false negative results in some cases are secondary to low viral loads likely associated with temporal aspects of viral shedding.

Our study indicates that prolonged viral RNA shedding is associated with recovery of infectious virus in a subset of patients and seems to correlate with persistence of symptoms. Higher Ct values and positive RNA tests detected after viral RNA clearance were not associated with recovery of infectious virus in our tested cohort. DdPCR can add an increased sensitivity in detecting viral RNA. Our data support the recently updated CDC guidelines for the duration of isolation after a positive COVID-19 test (23). Additional studies are required to inform using Ct values and cell culture results in making clinical decisions and developing diagnostic strategies that can differentiate shedding versus active replication will be very valuable for infection control.

## Data Availability

Virus sequence data has been submitted to public databases including GISAID and Genbank.

## Acknowledgements

We thank the entire clinical microbiology laboratory for the rapid response to the pandemic and for offering a unique testing capacity. This work was funded by the department of pathology, Johns Hopkins School of Medicine, the NIH Johns Hopkins Center of Excellence in Influenza Research and Surveillance HHSN272201400007C (A.P., H.H.M.) and T32A1007417 Molecular and Cellular Basis of Infectious Diseases (H.P. and B.S.). DdPCR was performed in collaboration with Bio-Rad Laboratories. We thank Winston Temp, PhD and Stuart C. Ray, MD for their valuable contribution to the SARS-CoV-2 genomic analysis pipeline.

